# Voxel-wise hemispheric Amyloid Asymmetry and its association with cerebral metabolism and grey matter density in cognitively normal older adults

**DOI:** 10.1101/2024.03.05.24303808

**Authors:** Hunsica J. Jayaprakash, Akiko Mizuno, Beth E. Snitz, Ann D. Cohen, Willian E. Klunk, Howard J. Aizenstein, Helmet T. Karim

## Abstract

**Introduction:** Alzheimer’s disease (AD) is a neurodegenerative disorder characterized by changes in beta amyloid (Aß) and tau as well as changes in cerebral glucose metabolism and gray matter volume. This has been categorized as three distinct stages of amyloid, tau, and neurodegeneration. Past studies have shown asymmetric Aβ accumulation and its association with asymmetric cerebral metabolism in preclinical AD. We analyzed data to replicate these findings and extend them to associations with gray matter volume and cognitive function.

**Methods:** We recruited 93 (mean age = 76.4±6.1 years) cognitively normal adults who underwent magnetic resonance imaging (MRI) and positron emission tomography (PET) with Pittsburgh compound B (PiB) and Fluorodeoxyglucose (FDG) tracers (to estimate Aβ and glucose metabolism, respectively). We conducted voxel-wise paired t-test on PiB (left vs. right hemispheres) to identify regions that differ in Aβ between the left and right cortex. We identified whether these regions showed asymmetry in FDG and gray matter volume using paired t-tests on each region. We then conducted correlations between asymmetry indices for each region that had significant asymmetry in PiB, FDG, and gray matter volume. We ran a group regression analysis on cognitive functions.

**Results:** We found 26 regions that had significant rightward asymmetry in PiB including prefrontal cortex, temporal cortex, insula, parahippocampus, caudate, and putamen. All these regions showed significant gray matter rightward asymmetry, and most of these regions showed significant FDG asymmetry except the caudate, orbital cortex, medial frontal gyrus, and superior temporal gyrus. Only in the superior frontal gyrus, we found that greater rightward asymmetry in PiB was associated with greater rightward asymmetry in FDG, *r*(82) =0.38, *p*<0.005 (FDR corrected) – no other regions showed significant Aß asymmetry correlation with either FDG or gray matter volume asymmetry. We found that greater rightward FDG asymmetry in the superior frontal gyrus was associated with greater visuospatial processing scores in our cognitive domain group regression analysis.

**Discussion:** AD has previously been modeled in three-stages: however, our results indicate that cerebral glucose metabolism may be dynamic throughout the disease progression and may serve as a compensatory pathway for maintaining cognitive functioning.

## Introduction

Alzheimer’s disease (AD) is the leading cause of dementia^1^ and has increased in prevalence due to an aging population^1^. While memory loss is a predominant symptom of AD, it also affects other cognitive domains including language, visuomotor processing, attention, and executive function^2^. Biomarkers of AD like beta amyloid (Aβ) can accumulate in various parts of the brain 15 years prior to AD symptoms^3^. In classical models of AD, this is followed by tau, neurodegeneration, and functional changes (e.g., cerebral glucose metabolism) and then eventually cognitive decline^4^. While Aβ accumulation occurs on average in certain patterns in the brain (e.g., predominantly in the medial temporal regions and posterior cingulate cortex^3,5,6^), there is significant Aβ deposition variability which may vary within individuals both temporally and spatially. This very early variability may drive early differences in neural and cognitive function. Specifically, there is evidence of asymmetric deposition of amyloid in patients with AD between the left and right hemisphere^7^.

In previous AD studies, significant Aβ asymmetry has been associated with asymmetric brain glucose metabolism (estimated by FDG PET)^7^. Greater Aß asymmetry may thus correlate with asymmetry in cerebral glucose metabolism, neurodegeneration, and even cognitive function. In the process of pathological aging and at later stages during AD progression, loss of functional or domain-specific specialization (known as dedifferentiation) occurs in which compensatory mechanisms emerge to maintain brain function^8,9^. Thus, to maintain cognitive function the brain becomes more domain general – recruiting resources more broadly and even bilaterally. However, not all asymmetry is associated with markers like Aß, structural and functional asymmetry can occur naturally in cognitively normal, healthy individuals. For example, language centers in the brain have an asymmetric distribution and are mostly known to be lateralized in the left hemisphere. While functional and structural asymmetry might be a naturally occurring phenomenon, later stages of AD demonstrate a unique relationship between Aβ and FDG.

There are few studies that have investigated the pattern of asymmetric (not limited to hemispheric but also focal) distribution of Aß. Several studies have shown that amyloid typically deposits within the default mode network and then frontal executive regions later^3,5,6^ though there is significant variability. One study in AD participants showed that different cognitive subtypes were associated with differing Aß deposition and cerebral glucose metabolism^7^. They found that Aß and cerebral glucose metabolism were greater on the right compared to the left hemisphere^7^ and these asymmetry between Aß and metabolism were associated in the angular gyrus, middle frontal gyrus, visual cortex, superior parietal gyrus, and middle and inferior temporal gyrus^7^ – showing hypometabolism on the side of greater Aß. They additionally showed that greater left (compared to right) hemisphere Aß was associated with more severe language impairment while greater right compared to left Aß was associated with more severe visuospatial impairment^7^. This result has been replicated in another study^10^. In the late stages of disease, greater Aß may drive lower cerebral metabolism – however earlier stages may show a reverse pattern (e.g., reflecting compensatory processes).

A recent study in participants without amyloid compared to those with amyloid compared to those with mild cognitive impairment (MCI) showed no asymmetry in those without amyloid, that there was greater asymmetry in the right hemisphere in the earliest stages, and again no asymmetry in late stages (i.e., in MCI)^11^. They found that these did not correlate with cognitive function^11^.

We sought to replicate the work of this past MCI study^11^ in cognitively normal older adults with varying Aß accumulation and conduct voxel-wise analyses of asymmetry which have not been done before. We also sought to extend these findings to cerebral metabolism and gray matter volume asymmetry as well as correlates with an extensive neurocognitive battery. We conducted voxel-wise analyses to evaluate left-right hemispheric differences in Aβ estimated by PiB and evaluated how these differences were related to asymmetry in cerebral glucose metabolism as well as gray matter volume, and finally their correlation with cognitive function.

## Methods

### Participants and Study Design

Data was obtained from an ongoing study at the Alzheimer’s Disease Research Center at the University of Pittsburgh (PI: Aizenstein & Villemagne, 2RF1AG 025516). We recruited cognitively normal older adults via advertisements or mailings to individuals who were interested in aging research. We analyzed all available data (n=93, mean age = 76.4±6.1 years) from participants who underwent scan sessions for MRI and positron PET with PiB and FDG tracers. The inclusion criteria were participants with normal cognitive function (see details below), and exclusion criteria included the presence of dementia or MCI (Mild Cognitive Impairment) history of major neurologic or psychiatric disease, Geriatric Depression Scale (30-item) greater than a clinical cutoff of 15, psychoactive medication use, contraindications to MRI, or having visual or auditory or motor deficits which may prevent the completion of behavioral testing.

Categorization of individuals as cognitively normal occurred if no more than two of their neuropsychological assessments were below one standard deviation below the age and education adjusted norms for that assessment. Detailed description of inclusion and exclusion criteria have been previously reported^12^. Participants gave written informed consent before enrolling. The University of Pittsburgh Institutional Review Board approved this.

### Cognitive Assessments

Each participant went through the battery of neurocognitive assessments, used by the University of Pittsburgh Alzheimer Disease Research Center (ADRC). The battery examined five domains of cognitive ability: memory learning, memory retrieval, visuospatial, language, and executive and attention. We used the following tests to access each domain: memory [Logical Memory^13^, Modified Rey-Osterreith Figure^14^, ADRC Word List^15^]; visuospatial construction ([Block Design, Modified Rey-Osterreith Figure Copy^16^)]; language ([Animal and Letter Fluency^17^, the 60-Item Boston Naming Test^18^]); and executive function ([Trail Making Test A and B^19^, Clock Drawing^20^, Maximum Digit Span Forward and Backward^13^]. We computed two memory domain scores by using the immediate and delayed recall scores for “memory learning” and “memory retrieval,” respectively.

### MRI Data Acquisition and Processing

Participants underwent 3T structural MRI scans using a 3T Siemens Trio TIM scanner with a 12-chanel head coil. Sagittal whole brain 3D magnetization prepared rapid-acquisition gradient echo (MPRAGE) was collected with echo time (TE)L=L2.98 msec, repetition time (TR)L=L2,300 msec, flip angle (FA)L=L9o, field of view (FOV)L=L256L×L240, 1L×L1L×L1.2 mm resolution, 0.6 mm gap, and GeneRalized Autocalibrating Partial Parallel Acquisition (GRAPPA) acceleration factorL=L2. Axial whole brain 2D fluid attenuated inversion recovery (FLAIR) was collected to measure WMH burden with TEL=L90 msec, TRL=L9,160 msec, FAL=L150o, FOV = 212L×L256, 1L×L1L×L3 mm resolution, no gap, and GRAPPAL=L2.

MRI then underwent standard preprocessing using Statistical Parametric Mapping (SPM12)^21^ toolbox in MATLAB2016b (Mathworks) and structural sequences were coregistered to MPRAGE, bias-corrected, and segmented into 6 classes^22^. Due to presence of white matter hyperintensities, we adjusted the number of Gaussians used to identify white matter to 2 to improve identification of gray and white matter^23^. We used Diffeomorphic Anatomical Registration using Exponentiated Lie Algebra (DARTEL) to generate a study template^24^. This approach uses each participant’s gray and white matter image to coregister to a standard anatomic space. Iterative average templates are generated and registered to, which completes once a final crisp average template is generated. This approach improves the normalization to a standard anatomic space. The gray matter was then transformed to this standard space and the total amount of gray matter is preserved by multiplying by the determinant of the Jacobian of the transformations.

### PET Data Acquisition and Processing

PET emission data were acquired on a Siemens ECAT HR+ PET scanners. Participants were positioned in the scanner approximately 35 min after [11C] PiB injection (15 mCi). Following the injection, a 10 min transmission scan was acquired using rotating 68Ge/68Ga rod sources were used to correct for photon attenuation, this was followed by a 20-minute emission scan (4 x 5-minute frames) beginning 50 minutes after [11C] PiB injection. FDG was injected intravenously (6-7.5mCi) and the PET scan was acquired over 25 min (five 5min frames) after a 35-minute uptake period where participants lay quietly in a dimly lit room with their eyes open. Filtered back-projection (Direct Inverse Fourier Transform) was used for reconstruction of PET emission data into a 128 x 128 x 63 matrix with voxel sizes of 2.06 x2.06 x 2.43 mm3. Images were then filtered with a 3 mm Hann window.

We inspected the dynamic PET (PiB and FDG separately) scans for interframe motion. If suspected, the automated image registration algorithm, optimized for PET-to-PET registration, was applied on a frame-wise basis. A summed image was generated over the post-injection interval. The FDG data were summed over the 40-60 min interval (4 frames). For both images, a standardized uptake value ratios (SUVR) were obtained by normalizing tissue radioactivity concentration (Ci/ml) by injected dose (mCi) and body mass (in units of ml, making the approximation that 1 g equals 1 ml) with respect to a reference region (i.e., the grey matter of the cerebellum). The average SUVR maps were registered to the MPRAGE in native space and then normalized to MNI space using the deformation field generated during structural processing.

### Statistical Analysis

We conducted statistical analysis on the demographic and clinical data in R (version 4.3.1). We conducted a voxel-wise paired t-test on PiB SUVR (left vs right hemispheres) using statistical non-parametric mapping toolbox (SnPM13)^25^. We did this by masking each image as left or right hemisphere and then flipping the right hemisphere images. SnPM computes non-parametric p-values using permutation testing (10,000 permutations). To adjust for multiple comparisons, we controlled the cluster-wise (uncorrected cluster forming threshold at *p*L<L0.001) family-wise error (FWE) rate at 0.05. For those regions that showed significant asymmetry, we extracted the averaged left and right hemisphere PiB SUVR of each region to compute an index of asymmetry (left minus right divided by the sum of left and right, i.e., greater positive values indicate greater left asymmetry). We separated clusters using the AAL3 atlas definitions for various regions^26^.

For each of those regions that showed significant PiB SUVR asymmetry, we also extracted left and right hemisphere FDG SUVR and gray matter volume. We conducted region-wise paired t-tests for the FDG SUVR and gray matter volume and adjusted for multiple comparisons using FDR correction (α=0.05). Then, for the regions that showed significant FDG SUVR and gray matter asymmetry, we computed an index of asymmetry of FDG SUVR and gray matter asymmetry, respectively.

In regions that showed significant FDG SUVR and gray matter volume asymmetry, we evaluated associations between those and PiB SUVR asymmetry. We conducted Pearson correlation analysis between the asymmetry indices of PiB SUVR and FDG SUVR as well as PiB SUVR and gray matter volume corrected for multiple comparisons using FDR (α=0.05).

We then conducted regression analysis for each cognitive domain (memory learning, memory retrieval, visuospatial, language, and executive and attention) as dependent variables and PiB SUVR asymmetry/FDG asymmetry/GMV asymmetry in any regions that were significant in all the tests above while adjusting for age, sex, race, education, and APOE. We conducted multiple imputations using the mice package in R and random forest method^27^.

## Results

The demographic and clinical characteristics of the sample are shown in Table 1. We identified 26 regions that showed significantly greater PiB SUVR on the right compared to the left hemisphere (but none in the opposite direction) – these are shown in Figure 1 and Table 2.

**Figure 1.**
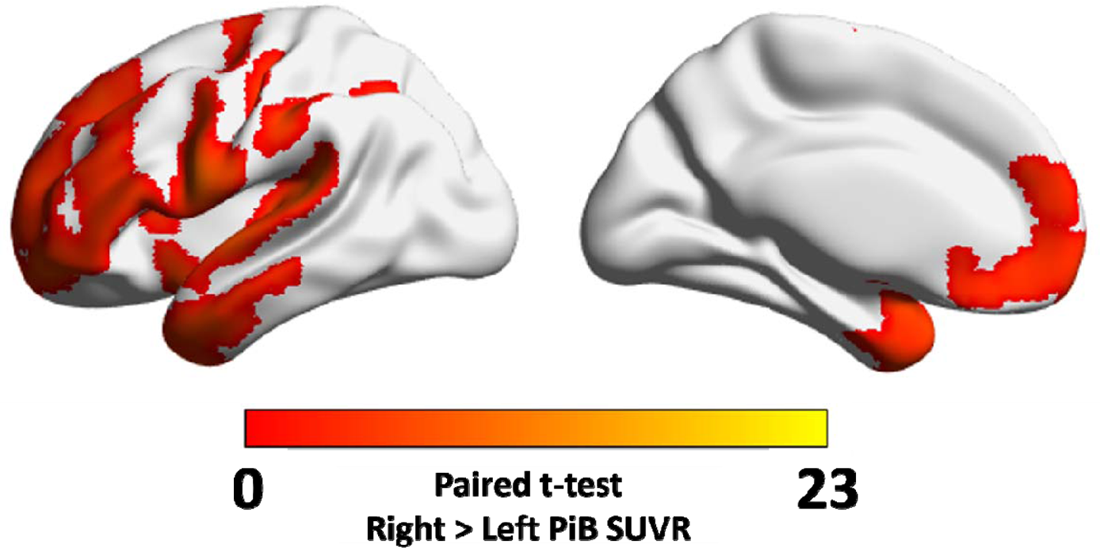
Voxel-wise differences between left and right PiB SUVR (paired t-test). All regions showed significantly greater PiB SUVR on the right compared to the left hemisphere. Note: these are all shown on a left hemisphere image as the right SUVR map was flipped onto the left for voxel-wise analysis. Images generated in BrainNetViewer.

**Table 1.** Demographic and clinical characteristics of the sample studied.

| Variables | Whole Group<br>(n=93) |
| --- | --- |
|  | mean (SD) or n (%) |
| age (years) | 75 (6) |
| Sex (male) | 27 (29%) |
| Race (non-White) | 11 (12%) |
| Education (years) | 15 (2) |
| Global PiB (SUVR) | 1.57 (0.54) |
| Global FDG (SUVR) | 1.63 (0.13) |
| WMH (volume, cubic mm) | 6728 (10182) |
| APOE (at least one E4 allele) | 18 (19%) |

**Table 2.** Regions that showed significantly greater PiB SUVR on the right compared to the left hemisphere. The cluster size (in voxels, 2mm^3^ resolution), max t-test value in that region, and the location of the max (in MNI space) are listed.

| Region | Cluster Size (voxels) | Paired T-test Max | MNI X | MNI Y | MNI Z |
| --- | --- | --- | --- | --- | --- |
| Superior Frontal | 2009 | 9.5 | -22 | 66 | 20 |
| Middle Frontal | 1748 | 14.6 | -50 | 40 | 20 |
| Superior Temporal | 1169 | 18.2 | -64 | -4 | -2 |
| Precentral Gyrus | 1134 | 14.4 | -60 | 10 | 30 |
| Inferior Frontal (Triangular) | 1078 | 14.3 | -56 | 20 | 4 |
| Superior Temporal Pole | 785 | 12.9 | -20 | 16 | -32 |
| Postcentral Gyrus | 772 | 14 | -64 | -2 | 22 |
| Middle Temporal | 699 | 16.7 | -64 | -8 | -16 |
| Medial Frontal (Orbital) | 666 | 6.9 | -4 | 68 | -16 |
| Insula | 614 | 8.6 | -34 | 6 | -16 |
| Middle Temporal Pole | 603 | 14 | -52 | 12 | -32 |
| Inferior Frontal (Oper) | 515 | 13.1 | -62 | 12 | 20 |
| Inferior Parietal | 497 | 7 | -50 | -30 | 40 |
| Putamen | 438 | 9.1 | -30 | -20 | 2 |
| Superior Medial Frontal | 437 | 6.1 | 0 | 68 | 2 |
| Inferior Temporal | 398 | 9.5 | -56 | -2 | -32 |
| Supramarginal | 360 | 13 | -64 | -24 | 34 |
| Rolandic Operculum | 312 | 13.5 | -64 | -4 | 4 |
| Pallidum | 282 | 8 | -24 | -14 | 2 |
| Fusiform Gyrus | 223 | 10.5 | -22 | 6 | -48 |
| Orbitofrontal (anterior) | 204 | 11.1 | -26 | 52 | -18 |
| Caudate | 201 | 6.2 | -14 | 6 | 10 |
| Heschl Gyrus | 179 | 12.5 | -64 | -10 | 8 |
| Parahippocampus | 158 | 5.1 | -20 | 0 | -26 |
| Inferior Frontal (Orbital) | 141 | 11 | -52 | 42 | -6 |
| Rectus Gyrus | 136 | 7.1 | -4 | 66 | -18 |

These regions included prefrontal cortex, medial and orbital frontal cortex, sensory/motor cortex, inferior parietal and supramarginal gyrus, temporal cortex, insula and parahippocampus, as well as parts of the basal ganglia.

We extracted the FDG SUVR for each region and found that 14 regions showed asymmetry (Supplemental Table 1): 10 leftward (left>right) asymmetries and 4 rightward. The following showed leftward (left greater than right) asymmetry: heschl gyrus, insula, rectus gyrus, and putamen. The following showed right greater than left asymmetry: middle frontal, superior frontal, inferior frontal (triangular and operculum), orbitofrontal (anterior), pre- and post-central gyrus, supramarginal gyrus, fusiform gyrus, inferior temporal, middle temporal and pole, superior temporal pole, and parahippocampus.

We extracted the gray matter volume for each region and found that all 26 regions showed significant asymmetry (Supplemental Table 2) in both directions. The following showed a left greater than right asymmetry: middle frontal, postcentral gyrus, supramarginal, parahippocampus, insula, middle temporal, inferior temporal, fusiform gyrus, putamen, caudate, and pallidum. The following showed right greater than left asymmetry: superior frontal, inferior frontal, medial frontal, orbitofrontal (anterior), superior medial frontal, rectus gyrus, precentral gyrus and rolandic operculum, inferior parietal, superior temporal gyrus and pole, middle temporal, and heschl gyrus.

In each region that showed both significant PiB asymmetry and FDG asymmetry, we conducted correlations between those measures and found that greater rightward PiB asymmetry was associated with greater rightward FDG asymmetry in the superior frontal gyrus (Broadman Area 10) [see Figure 2, *r*(81)=0.38, *p*<0.001, *q*<0.01], but no other associations. In each region that showed both significant PiB asymmetry and gray matter asymmetry, we conducted correlations between those measures but found no significant association after adjusting for multiple comparisons.

**Figure 2.**
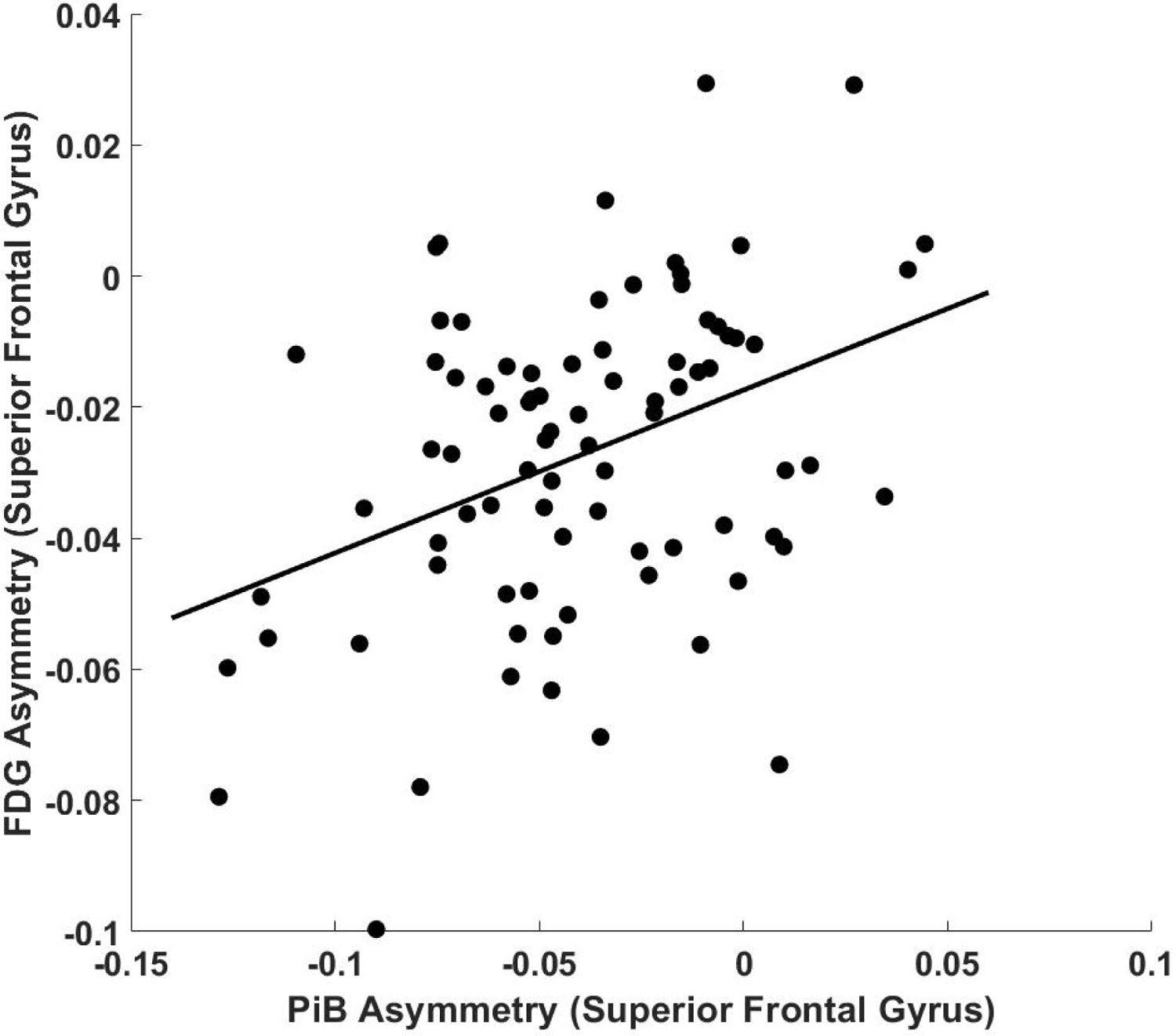
Correlation between superior frontal gyrus PiB asymmetry and FDG asymmetry. Positive asymmetry values indicate left greater than right asymmetry, while a value of zero indicates no asymmetry. Greater right PiB asymmetry was associated with greater right FDG asymmetry in the superior frontal gyrus.

We conducted regressions for each cognitive domain as a dependent variable and the following independent variables: superior frontal gyrus PiB and FDG asymmetry while adjusting for age, sex, Race, education, and APOE status. We found that greater rightward FDG asymmetry (i.e., greater FDG asymmetry index) was associated with better visuospatial function (Table 3, Figure 3) – we found no other associations with other domains (not shown).

**Figure 3.**
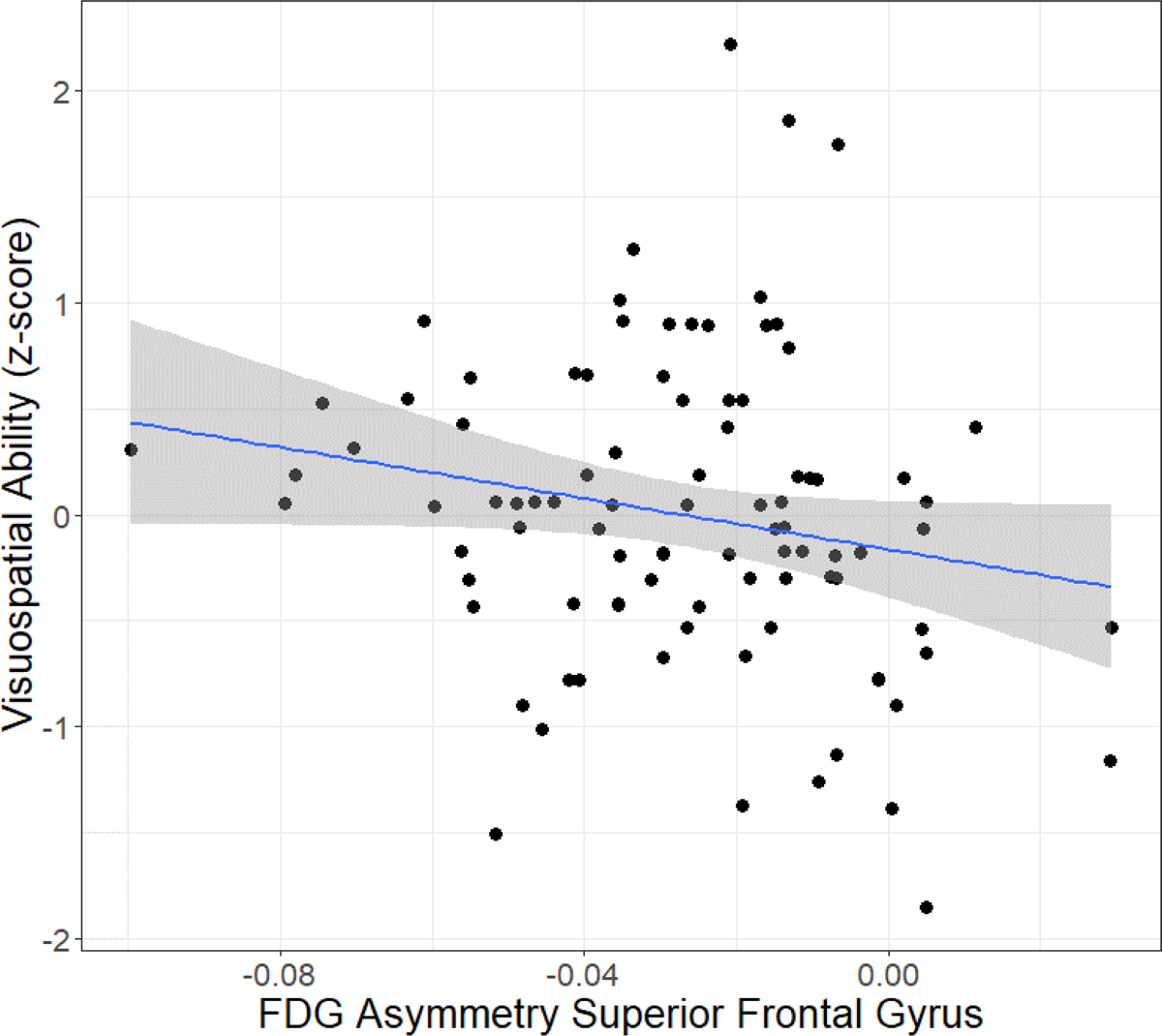
Plot between FDG asymmetry in the superior frontal gyrus (arbitrary units) and visuospatial ability (z-score average of multiple cognitive assessments, see methods). Greater right asymmetry in FDG superior frontal gyrus was associated with better visuospatial ability.

**Table 3.** Regression between visuospatial cognitive domain and superior frontal gyrus PiB and FDG asymmetry while adjusting for age, sex, Race, education, and APOE status.

| Measure | B (std. error) | t-value |
| --- | --- | --- |
| Age | -0.029 (0.013) | -2.3* |
| Sex (Ref: Male) | -0.044 (0.161) | -0.3 |
| Race (Ref: White) | 0.115 (0.217) | 0.5 |
| Education | 0.059 (0.033) | 1.8 |
| APOE (Ref: No E4 Allele) | -0.108 (0.18) | -0.6 |
| Superior Frontal PiB asymmetry | 0.878 (2.175) | 0.4 |
| Superior Frontal FDG asymmetry | -6.496 (3.365) | -1.9* |

## Discussion

We identified significant rightward (right > left) asymmetry in Aß deposition in 26 regions primarily in the prefrontal cortex and the anterior parts of the default mode network. A number of these (14 out of 26) regions showed significant asymmetry in glucose metabolism (i.e., FDG SUVR) and all 26 showed significant gray matter volume asymmetry (both rightward directions); however, only one region showed significant association between Aß and FDG asymmetry which was the superior frontal gyrus (Brodmann Area 10). Greater rightward Aß asymmetry was associated with greater rightward FDG asymmetry, which was in turn correlated with greater visuospatial cognitive function. This replicates a recent study that showed that Aß deposited asymmetrically (right greater than left) in the earliest stages of AD (cognitively normal with no or low amyloid to preclinical to MCI)^11^ but then deposits symmetrically in both hemispheres in the later stages (i.e., MCI). Our results extended their findings to the association with cognitive function as well as corresponding rightward glucose metabolism asymmetry – a potential compensatory mechanism. The observed associations in this study may be an early functional sign that there is hypermetabolism in this region potentially due to underlying accumulation of amyloid to help maintain cognitive function, especially within the visuospatial domain.

These results replicate one past study in preclinical AD that showed higher rightward asymmetry with greater levels of amyloid but no asymmetry prior to any amyloid accumulation^11^. We found that there was right-hemisphere Aß asymmetry voxel-wise, whereas this previous study^11^ found that there was greater asymmetry in general correlated with Aß in a quadratic association peaking in those with heightened amyloid. Our study primarily focused on people with no cognitive impairment but included individuals with significant levels of Aß. The current analysis also conducted voxel-wise analysis to investigate differences in Aß, whereas Kjeldsen et al^11^ investigated Aß differences in regions of interest and asymmetry. Thus, our results extend these findings to include a number of regions that show early rightward asymmetry in Aß, but also asymmetry in FDG and gray matter volume. These results are also in contrast to another study that evaluated regional asymmetry, where they found that PiB was fairly symmetric in a smaller sample across diagnoses but found left greater than right PiB asymmetry in the dorsal frontal cortex and sensory motor area but the reverse pattern in the occipital cortex^28^. This study evaluated 17 participants some of who had MCI and AD – our results extend this to a much larger sample of cognitively healthy participants.

These results also extend findings to include that we observed a significant association between greater right FDG asymmetry and greater right Aß asymmetry in cognitively normal older adults. One past study with AD patients has shown that Aß asymmetry is correlated with cerebral glucose metabolism asymmetry^7^ but inversely – a potential indicator of disruption of neural function. At first this finding seems to contradict the current findings; however, we may expect that in the early stages there may be a compensatory increase in cerebral metabolism that ‘fails’ in later stages – this tracks well with the findings in early vs. later stages.

We also evaluated how these measures of asymmetry correlated with cognitive function. We found that the right FDG asymmetry was associated with higher visuospatial function. This result does not completely align with the previous findings. Although the study in AD participants showed that rightward asymmetry in Aß (not glucose metabolism) was associated with visuospatial dysfunction^7^, another study which included those with and without AD did not find any association between cognitive function and Aß asymmetry^11^. In our study, we also did not find a direction association of Aß asymmetry with cognitive function in any domain. We did find that asymmetry of glucose metabolism, however, was correlated with visuospatial function – potentially serving as a compensatory process. These early functional processes may be responsible for maintaining cognitive function in the presence of Aß. Whereas in later stages of AD, we can hypothesize that Aß severity may be associated with now failing functional systems (e.g., lower cerebral glucose metabolism) and thus worse cognitive performance. This is particularly interesting because there is new evidence that some of the earliest and most sensitive cognitive changes are within visuospatial function^29^, where one visuospatial test (card rotations) showed an inflection point in cognitive function 15 years prior to AD onset.

We did not find associations between Aß and gray matter volume asymmetry, which is not necessarily surprising as in cognitively healthy individuals. We expected fairly low neurodegenerative processes (though there may be some). In line with the previous hypothesis, we may expect gray matter asymmetry may be associated with failing hypometabolism in later stages. In the current study, we highlight that many of these regions showed gray matter volume asymmetry but no correlation with Aß asymmetry.

There are several limitations in this study. We analyzed data cross-sectionally, thus causal statements cannot be made. Our sample size consisted of cognitively normal individuals whose age range was fairly limited; thus, it is unclear how this relates to participants with MCI or AD or even older adults. We focused on PiB asymmetry and its association with FDG and gray matter volume asymmetry; however, the opposite effect may also exist. This is a study focused on cognitively healthy older adults and as such has some Aß positive individuals. We did not measure tau deposition in this sample, which is a critical factor here though this is in the earliest stages of disease.

The current study replicates and expands past results on Aß asymmetry. We found that Aß is greater in the right compared to left hemisphere and that similar regions also show both cerebral glucose metabolism and gray matter volumetric asymmetry, though only the superior frontal gyrus showed an association between right Aß asymmetry and right cerebral glucose metabolism asymmetry. Asymmetry in the superior frontal gyrus glucose metabolism correlated with visuospatial function. These results demonstrate early asymmetry in Aß may drive both neurofunctional and cognitive differences.

## Data Availability

Data is available upon request.

---

**Supplemental Table 1.** Regions that showed significant asymmetry in gray matter volume. Bolded region names showed significant asymmetry between left and right hemisphere. Positive values indicate that the left hemisphere has greater volume than the right hemisphere. Corrected for multiple comparisons correction using FDR (see q-values).

| Region Name | Paired T-test Left vs. Right (df=82) | p-value | q-value |
| --- | --- | --- | --- |
| Caudate | 39.5 | 0.0000 | 0.0000 |
| Inferior Frontal (Operculum) | -28.4 | 0.0000 | 0.0000 |
| Inferior Frontal (Orbital) | -23.6 | 0.0000 | 0.0000 |
| Inferior Frontal (Triangular) | -6.5 | 0.0000 | 0.0000 |
| Medial Frontal (Orbital) | -2.9 | 0.0049 | 0.0049 |
| Middle Frontal | 17.3 | 0.0000 | 0.0000 |
| Superior Frontal | -16.8 | 0.0000 | 0.0000 |
| Superior Medial Frontal | -33.1 | 0.0000 | 0.0000 |
| Fusiform Gyrus | 11.4 | 0.0000 | 0.0000 |
| Heschl Gyrus | -4.5 | 0.0000 | 0.0000 |
| Insula | 12.0 | 0.0000 | 0.0000 |
| Orbitofrontal (anterior) | -9.5 | 0.0000 | 0.0000 |
| Pallidum | 22.3 | 0.0000 | 0.0000 |
| Parahippocampus | 17.2 | 0.0000 | 0.0000 |
| Inferior Parietal | -41.3 | 0.0000 | 0.0000 |
| Postcentral Gyrus | 32.9 | 0.0000 | 0.0000 |
| Precentral Gyrus | -57.2 | 0.0000 | 0.0000 |
| Putamen | 4.7 | 0.0000 | 0.0000 |
| Rectus Gyrus | -9.9 | 0.0000 | 0.0000 |
| Rolandic Operculum | -3.2 | 0.0021 | 0.0022 |
| Supramarginal Gyrus | 24.2 | 0.0000 | 0.0000 |
| Inferior Temporal | 17.5 | 0.0000 | 0.0000 |
| Middle Temporal | 28.9 | 0.0000 | 0.0000 |
| Middle Temporal Pole | -24.3 | 0.0000 | 0.0000 |
| Superior Temporal Pole | -34.1 | 0.0000 | 0.0000 |
| Superior Temporal | -16.4 | 0.0000 | 0.0000 |

**Supplemental Table 2.**
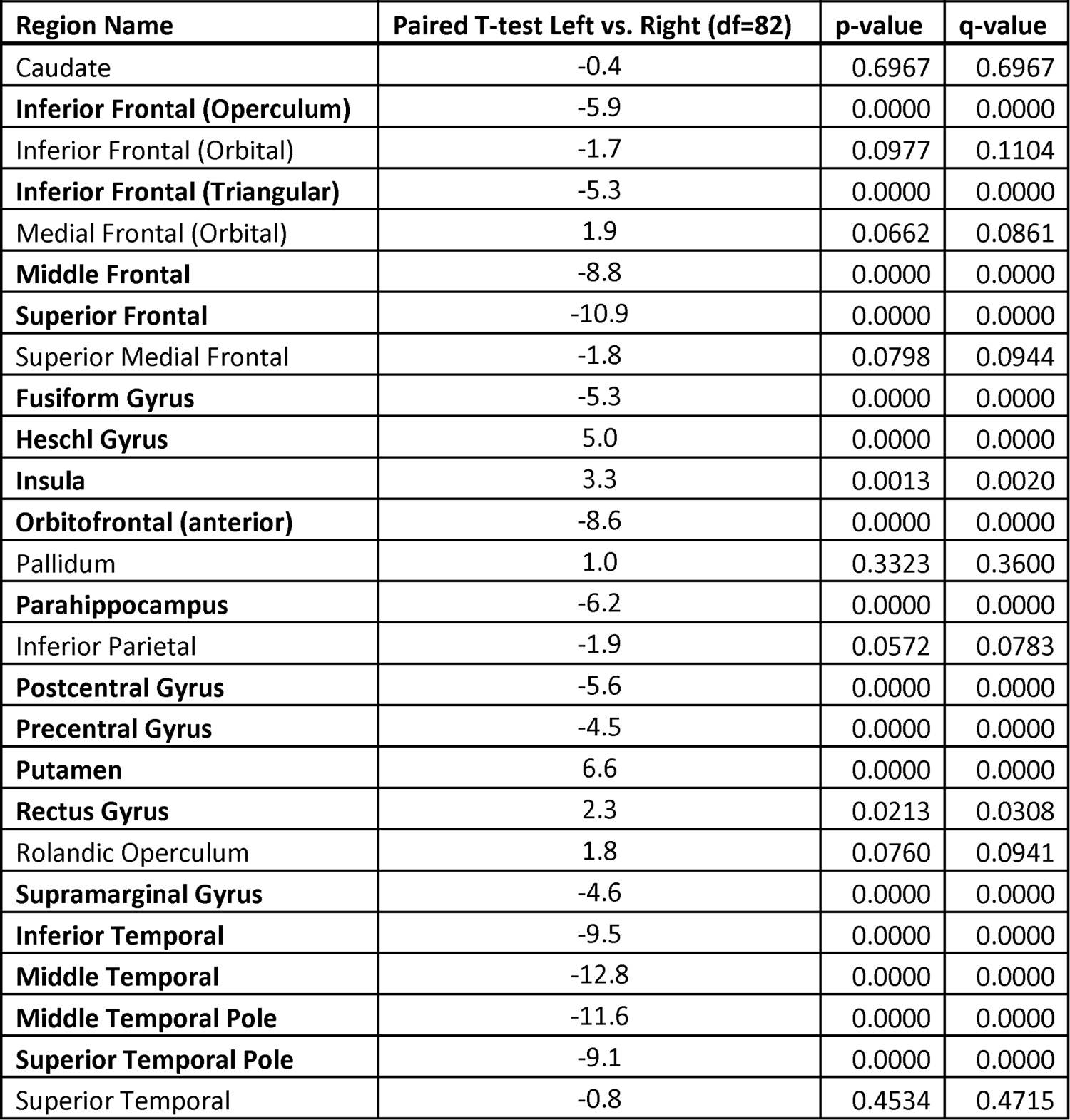
Regions that showed significant asymmetry in FDG. Bolded region names showed significant asymmetry between left and right hemisphere. Positive values indicate that the left hemisphere has greater FDG than the right hemisphere. Corrected for multiple comparisons correction using FDR (see q-values).

## Notes

### Competing Interest Statement

GE Healthcare holds a license agreement with the University of Pittsburgh based on the technology described in this manuscript. Dr. Klunk is a co-inventor of PiB and, as such, has a financial interest in this license agreement. GE Healthcare provided no grant support for this study and had no role in the design or interpretation of results or preparation of the manuscript. The other authors declare that they no competing interests.

### Funding Statement

This study was supported by P50 AG005133, P01 AG025204, and R37 AG025516 from the National Institute of Health.

### Author Declarations

Detailed description of inclusion and exclusion criteria have been previously reported. Participants gave written informed consent before enrolling. The University of Pittsburgh Institutional Review Board approved this.

## References

1. 2022 Alzheimer’s disease facts and figures. Alzheimers Dement. Apr 2022;18(4):700–789. doi:10.1002/alz.12638

2. Backman L, Jones S, Berger AK, Laukka EJ, Small BJ. Cognitive impairment in preclinical Alzheimer’s disease: a meta-analysis. Neuropsychology. Jul 2005;19(4):520–31. doi:10.1037/0894-4105.19.4.520

3. Palmqvist S, Scholl M, Strandberg O, et al. Earliest accumulation of beta-amyloid occurs within the default-mode network and concurrently affects brain connectivity. Nat Commun. Oct 31 2017;8(1):1214. doi:10.1038/s41467-017-01150-x

4. Jack CR, Jr., Knopman DS, Jagust WJ, et al. Tracking pathophysiological processes in Alzheimer’s disease: an updated hypothetical model of dynamic biomarkers. Lancet Neurol. Feb 2013;12(2):207–16. doi:10.1016/S1474-4422(12)70291-0

5. Grothe MJ, Barthel H, Sepulcre J, et al. In vivo staging of regional amyloid deposition. Neurology. Nov 14 2017;89(20):2031–2038. doi:10.1212/WNL.0000000000004643

6. Thal DR, Rub U, Orantes M, Braak H. Phases of A beta-deposition in the human brain and its relevance for the development of AD. Neurology. Jun 25 2002;58(12):1791–800. doi:10.1212/wnl.58.12.1791

7. Frings L, Hellwig S, Spehl TS, et al. Asymmetries of amyloid-beta burden and neuronal dysfunction are positively correlated in Alzheimer’s disease. Brain. Oct 2015;138(Pt 10):3089–99. doi:10.1093/brain/awv229

8. Park DC, Polk TA, Park R, Minear M, Savage A, Smith MR. Aging reduces neural specialization in ventral visual cortex. Proc Natl Acad Sci U S A. Aug 31 2004;101(35):13091–5. doi:10.1073/pnas.0405148101

9. Cabeza R. Hemispheric asymmetry reduction in older adults: the HAROLD model. Psychol Aging. Mar 2002;17(1):85–100. doi:10.1037//0882-7974.17.1.85

10. Yoon HJ, Kim BS, Jeong JH, Kim GH, Park HK, Chun MY. Asymmetric Amyloid Deposition as an Early Sign of Progression in Mild Cognitive Impairment Due to Alzheimer Disease. Clin Nucl Med. Jul 1 2021;46(7):527–531. doi:10.1097/RLU.0000000000003662

11. Kjeldsen PL, Parbo P, Hansen KV, et al. Asymmetric amyloid deposition in preclinical Alzheimer’s disease: A PET study. Aging Brain. 2022;2:100048. doi:10.1016/j.nbas.2022.100048

12. Snitz BE, Weissfeld LA, Cohen AD, et al. Subjective Cognitive Complaints, Personality and Brain Amyloid-beta in Cognitively Normal Older Adults. Am J Geriatr Psychiatry. Sep 2015;23(9):985–93. doi:10.1016/j.jagp.2015.01.008

13. Elwood RW. The Wechsler Memory Scale-Revised: psychometric characteristics and clinical application. Neuropsychol Rev. Jun 1991;2(2):179–201. doi:10.1007/BF01109053

14. Becker JT, Boller F, Saxton J, McGonigle-Gibson KL. Normal rates of forgetting of verbal and non-verbal material in Alzheimer’s disease. Cortex. Mar 1987;23(1):59–72. doi:10.1016/s0010-9452(87)80019-9

15. Morris JC, Heyman A, Mohs RC, et al. The Consortium to Establish a Registry for Alzheimer’s Disease (CERAD). Part I. Clinical and neuropsychological assessment of Alzheimer’s disease. Neurology. Sep 1989;39(9):1159–65. doi:10.1212/wnl.39.9.1159

16. Wechsler D. WAIS-R manual: Wechsler adult intelligence scale-revised. Psychological Corporation; 1981.

17. Strauss E, Sherman EM, Spreen O. A compendium of neuropsychological tests: Administration, norms, and commentary. American chemical society; 2006.

18. Goodglass H, Kaplan E, Weintraub S. Boston naming test. Lea & Febiger Philadelphia, PA; 1983.

19. Reitan RM. Validity of the Trail Making Test as an indicator of organic brain damage. Perceptual and motor skills. 1958;8(3):271–276.

20. Freedman M, Leach L, Kaplan E, Shulman K, Delis DC. Clock drawing: A neuropsychological analysis. Oxford University Press, USA; 1994.

21. Penny WD, Friston KJ, Ashburner JT, Kiebel SJ, Nichols TE. Statistical parametric mapping: the analysis of functional brain images. Elsevier; 2011.

22. Ashburner J, Friston KJ. Unified segmentation. Neuroimage. Jul 1 2005;26(3):839–51. doi:10.1016/j.neuroimage.2005.02.018

23. Karim HT, Andreescu C, MacCloud RL, et al. The effects of white matter disease on the accuracy of automated segmentation. Psychiatry Res Neuroimaging. Jul 30 2016;253:7–14. doi:10.1016/j.pscychresns.2016.05.003

24. Ashburner J. A fast diffeomorphic image registration algorithm. Neuroimage. Oct 15 2007;38(1):95–113. doi:10.1016/j.neuroimage.2007.07.007

25. Nichols TE, Holmes AP. Nonparametric permutation tests for functional neuroimaging: a primer with examples. Hum Brain Mapp. Jan 2002;15(1):1–25. doi:10.1002/hbm.1058

26. Rolls ET, Huang CC, Lin CP, Feng J, Joliot M. Automated anatomical labelling atlas 3. Neuroimage. Feb 1 2020;206:116189. doi:10.1016/j.neuroimage.2019.116189

27. Van Buuren S, Groothuis-Oudshoorn K. mice: Multivariate imputation by chained equations in R. Journal of statistical software. 2011;45:1–67.

28. Raji CA, Becker JT, Tsopelas ND, et al. Characterizing regional correlation, laterality and symmetry of amyloid deposition in mild cognitive impairment and Alzheimer’s disease with Pittsburgh Compound B. J Neurosci Methods. Jul 30 2008;172(2):277–82. doi:10.1016/j.jneumeth.2008.05.005

29. Williams OA, An Y, Armstrong NM, Kitner-Triolo M, Ferrucci L, Resnick SM. Profiles of Cognitive Change in Preclinical and Prodromal Alzheimer’s Disease Using Change-Point Analysis. J Alzheimers Dis. 2020;75(4):1169–1180. doi:10.3233/JAD-191268

